# When Diagnosis Is Made By Playing “Shuffled Cards”: A Non-Invasive, Real-Time, Adaptive Method to Functionally Evaluate Coronary Artery Based on Coronary Angiography

**DOI:** 10.1101/2022.03.01.22271738

**Authors:** Yanan Dai, Pengxiong Zhu, Bangde Xue, Yun Ling, Xibao Shi, Liang Geng, Yunhao Xie, Qi Zhang, Jun Liu

**Author notes:** Corresponding authors. Correspondence Shanghai East Hospital, Tongji University. No.150, Jimo Road, Shanghai, China Email address. The authors contributed equally to this work (Yanan Dai, Pengxiong Zhu).

## Abstract

**Aims:** The great value concealed in the sequence of coronary angiography frames is not discovered in the world. We discovered and demonstrated the “Sequence Value” concealed in the coronary angiography and proposed DZL (Disarranged Zone Learning) to realize the sequence value in functional evaluation of coronary artery disease. Furthermore, we automated the DZL using a deep learning model to release huge medical resources.

**Methods and Results:** We gave a novel definition of TIMI flow grade using the term temporal and spatial coupling tightness (TSCT) of the antegrade contrast agent. We used the TSCT to model the myocardial ischemia in a functional perspective and used the PCI conduction after CAG as the outcome event of myocardial ischemia. We proposed a novel method (Disarranged Zone Learning) to measure TSCT and we designed an experiment to validate its effectiveness. We further automated the novel method using an unsupervised deep learning model. The prediction accuracy of the model was applied as a proxy of myocardial ischemia. We further proposed Difference DZL to quantify the functional capability of any specific vessel segment. DZL overall AUC reaches 0.92. DZL automation reveals an AUC of 0.84 (95%CI, 0.81-0.87).

**Conclusion:** We unprecedentedly discovered the “Sequence Value” concealed in coronary angiography. We then proposed a novel method termed DZL to functionally evaluate the coronary artery in a non-invasive, real-time and adaptive manner.

**Disclosure:** The Authors declare that there is no conflict of interest.

## 1. Introduction

Hemodynamics of the coronary artery system is a complicated fluid dynamics mechanism. It is affected by many factors such as vasodilatation, blood pressure, heart rate, artery anatomic structure (lumen diameters, vessel lengths, branches, bending angles etc.), blood viscosity and density etc. ^1^ Recent studies have demonstrated that myocardial ischemia is not sufficiently indicated merely by artery anatomic structure, conventionally mapped to artery stenosis, but rather comprehensively on a functional perspective intrinsically rooted in hemodynamics. ^2^ However, before FFR being regarded as a standard and effective approach to discriminate stenosis causing pathological myocardial ischemia while exclude stenosis with “wolf clothing but sheep inside”, CAG is recognized as the gold rule to diagnose coronary artery disease in an anatomic perspective. Unfortunately, anatomic stenosis does not necessarily mean functional disability such as myocardial ischemia. CAG is not a functional diagnosis approach, some important functional features fail to be extracted and are blinded, causing the false positive and false negative diagnosis. False positive diagnosis leads to unnecessary procedure and false negative diagnosis leads to improper delayed treatment. FFR was thus developed and able to evaluate coronary artery functionally. ^3^ However, although the consensus is reached that FFR is the gold rule to deliver precise functional diagnosis of the coronary artery, it is an invasive procedure bringing extra operation durations, risks, expenses. All these disadvantages of FFR prevent it from general clinical application. To relief the dilemma, recent study has demonstrated the possibility and effectiveness of QFR to non-invasively calculate a proxy FFR using coronary angiography. ^4^ However, QFR belongs to a simulation method, it simulated the hemodynamics based on fluid dynamics. Such prior knowledge relied method faces challenges in adaptive capability. In case that the blood vessels tree is physically restructured or repaired so that the prior regularizations are violated, the effectiveness of the method is degenerated. For example, patients with cardiac surgery or operation history will be the outliers to have limited effectiveness of QFR. Besides, QFR is less able to evaluate prognostic effectiveness right after the PCI since the property of the stent placed and the physiological reactions successively are out of the scope of the simulation algorithm. Lastly, the necessary condition for QFR algorithm to work is the input of two coronary angiography images of different angles projecting the same vessel and the angle difference is at least 25.^5^ Such hard constraint restrains parts of coronary angiography from being re-evaluated ex post. In order to make ex post evaluation, even for the coronary angiography with qualified angles, manual efforts are required to select the suitable images of different angles as the input to the QFR algorithm which is laborious.

In this study, we uncovered the “Sequence Value” concealed in coronary angiography and proposed a novel method to measure functional capability of coronary artery based on coronary angiography. Even for patients with previous cardiac surgeries or operations, the method is applicable as well since it is not a simulation and remodeling based method but able to capture the true fluid dynamics, the adaptiveness of the novel method is strong. Besides, the method gains the ability to guide revascularization procedure in the real-time perspective for it works within dozens of seconds. Furthermore, it is able to detect instant post-revascularization effectiveness during the procedure in real-time. As a byproduct, such method facilitates the evaluation of huge amounts of history coronary angiographies in a functional way, pushing the retrospective medical study forward heavily since the history data is able to be re-evaluated in mass production without manual efforts involved. Lastly, we automated the method using an unsupervised deep learning model to liberate the huge medical resources and enable the large real-life clinical applications.

## 2. Methods

The method is termed Disarranged Zone Learning. The overall framework of DZL is illustrated in Figure 1. Firstly, we introduced a novel definition of TIMI flow grade using TSCT. Then we uncovered the relationship between TSCT and Sequence Recovery Capability and validated the corresponding link between Myocardial Ischemia and Sequence Recovery Capability using an experiment. Later, we defined ischemia score and set PCI conduction after CAG as the outcome event. Lastly, we developed an unsupervised deep learning model to automate DZL.

**Figure 1.**
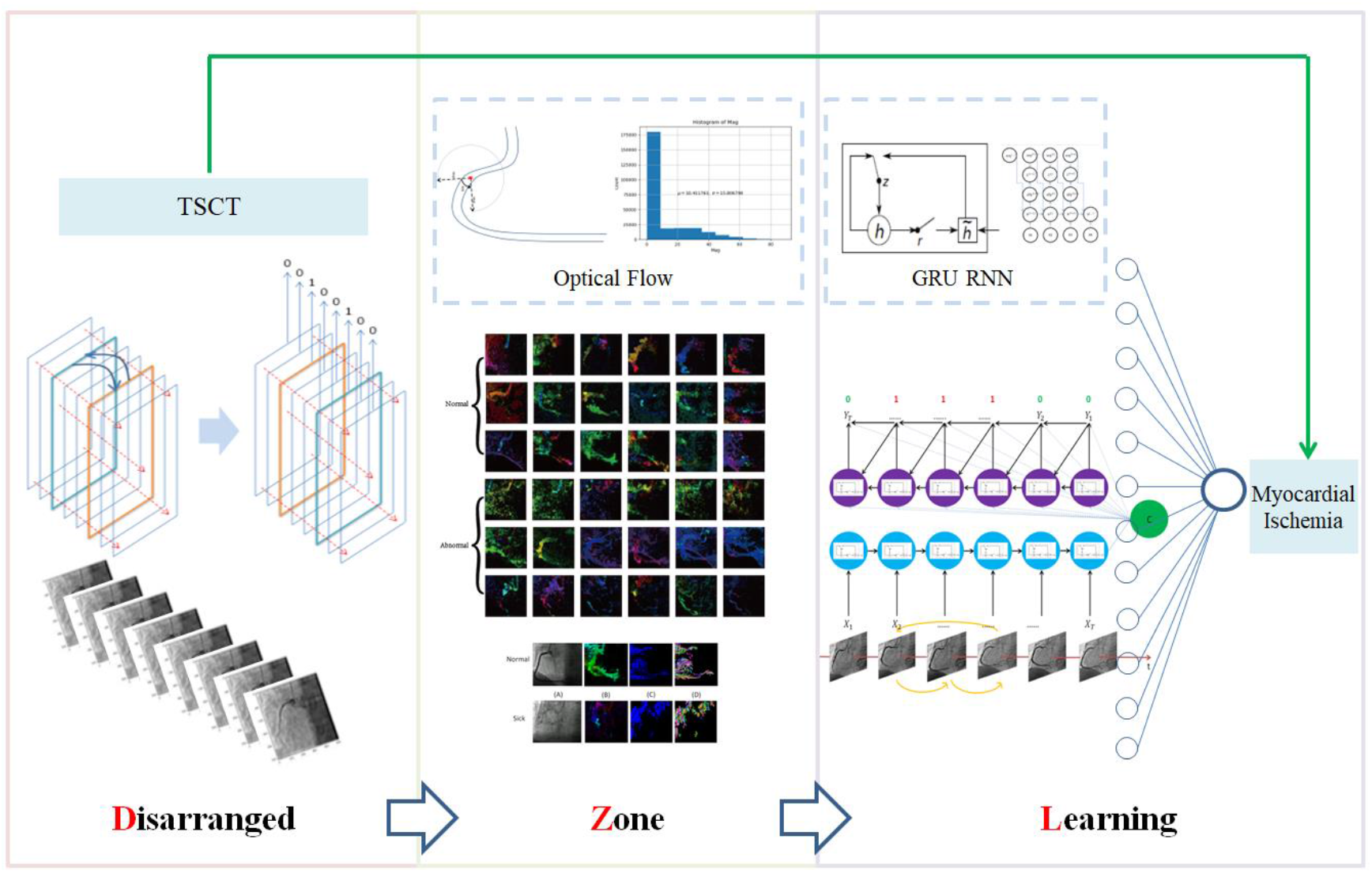
Overall Framework. The overall method can be divided into three parts, including the disarranged, zone and learning. The disarranged part is functional to disorder the frames of angiography video. We shuffled half of the total frames in a video in order to keep the balance of positive and negative input. The zone part is functional to generate an effective zone to capture the sequence information of video. For DZL automation, we use optical flow to reduce the data dimension and generate sequential representations of the input. The learning part is to learn how to recover the disarranged order accordingly. For DZL automation, an Encoder Decoder GRU neural network is designed and applied to act as the core of the unsupervised model.

### 2.1 Model Coronary Angiography in a Functional Perspective: TIMI flow grade

Coronary angiography (CAG) is exhibited in a form of video, which records the dynamic process of contrast agent flowing in the blood vessel. The part that is unable to be developed by contrast agent is the location of suspected lesions. ^6^ Investigation on the way to reflect the lesions causes the analysis of coronary angiography extended to the three-dimensional (3D) space, by adding the extra dimension of time. The strong dependence makes the analysis of coronary angiography dependent on the whole of the video. The correlation between the adjacent images makes the frame sequence unable to maintain Markov property ^7^, hence further increase the complexity of modeling.

We modeled the coronary angiography in a functional TIMI flow grade perspective. From the definition of TIMI flow grade ^8^, we proposed a novel equivalent TIMI flow grade definition based on temporal and spatial coupling tightness of the antegrade contrast agent (TSCT). Based on the novel definition, the higher the TSCT is, the higher the TIMI flow grade is. Then we later proposed a novel method to measure the TSCT. There are two branches of theoretical arguments in terms of how human brains process temporal and spatial information: CMT (Conceptual Metaphor Theory) v.s. ATOM (A Theory of Magnitudes). CMT is proposed by Lakoff and Johnson ^9^ which assumes that the neural system characterizing concrete sensorimotor experience has more inferential connections and therefore a greater inferential capacity than the neural system characterizing abstract thoughts. ^10^ The abstract expression of time is thus dominated by the concrete expression of space, forming an asymmetric hypothesis. Time can be expressed by space more than space expressed by time. ^11^ The ATOM is proposed by Walsh ^12^ proposing hypothesis that time and space information are processed physically activating overlapped area of the brain (parietal cortex), which indicates that there exists an unified magnitude for humans to process information in these two separating domains. Thus, ATOM is featured as the symmetric hypothesis, temporal and spatial information weight equally and can be exchangeable as the processing output signal to activate further action. ^12^ Whichever theory we choose to build on, it supports the argument that time can be represented by space, this is the foundation rooted on our novel method to measure the coupling tightness between time and space.

### 2.2 A Novel TIMI (Thrombolysis In Myocardial Infarction) Flow Grade Definition

TIMI flow grade is divided into 4 grades: no perfusion, penetration without perfusion, partial perfusion, complete perfusion.^8^ Recent studies demonstrated that obvious disparities between stratified TIMI grades of these two groups (TIMI 3 v.s.TIMI ≤2) ^13^. For the group of TIMI3, superior clinical outcome such as global left ventricular function, lower enzyme peaks, reduced morbidity and mortality rates is demonstrated.

TIMI flow grade is evaluated depending on perfusion extent and indicated by opacified rate. The opacification process, in a human visual perspective, is a temporal and spatial integrative physical movement. The antegrade contrast material moves forward in a specified time window and the process is evaluated to generate the TIMI flow grade. The spatial movement alone is not sufficient to grade as the temporal constraint is required to be imposed to distinguish Grade2 and Grade3 since the complete perfusion is defined that the antegrade contrast material should enter or clear(or both) with predefined speed. Brain imaging studies consistently show that time, space, number and other magnitudes such as size and brightness activate overlapping regions in the brain part termed parietal cortex.^12^ This is also the physical foundation of ATOM. The interactive temporal and spatial indications of antegrade contrast material activate the area of parietal cortex and enable the generation of TIMI flow grade by human brain. Consequently, based on the original TIMI flow grade definitions^8^, we proposed the following novel definitions of TIMI grade flow accordingly using TSCT.

- Grade0: the temporal and spatial contrast material indications of the obstruction are completely decoupled. With the time goes on, no antegrade contrast material is beyond the area of obstruction and thus no spatial movement of contrast material is detected.
- Grade1: the temporal and spatial contrast material indications of the obstruction are nearly decouples. Only within a short period while the antegrade contrast material passing the obstruction that the spatial movement of contrast material is detected. After the time period, as the contrast material failed to reach coronary bed distal, with the time goes on, no antegrade contrast material is observed and no spatial movement of contrast material is detected.
- Grade2: the temporal and spatial contrast material indications of the obstruction are inefficiently coupled. With the time goes on, corresponding antegrade contrast material is detected to reach the coronary bed distal. However, the whole process works slowly and inefficiently comparing to other areas not perfused by the previous occluded vessel.
- Grade3: the temporal and spatial contrast material indications of the obstruction are efficiently coupled. With the time goes on, corresponding antegrade contrast material is detected and reach the coronary bed distal efficiently.

### 2.3 The Insight Behind Sequence Recovery Capability: A TIMI Flow Grade Measurement

From the above definition, it is elucidated that the tighter antegrade contrast agent is coupled temporally and spatially, the higher TIMI flow grade it is. Following this logic, for the tightly temporally and spatially coupled antegrade contrast agent, if we shuffle the time factor, it will thus rearrange the antegrade contrast agent spatially. The approach we implemented to shuffle the time factor is to shuffle the coronary angiography frame sequences and form a new disordered film. Since the disordered time will cause spatial incoherency of opacification process of contrast agent and such incoherency, if reaches some extent, is possible to be observed by humans visually and also be eliminated through sequence recovery. On the contrary, if the shuffled time brings no obvious spatial incoherency, and humans are not able to distinguish the shuffled frames, let alone to recover the sequence. According to such arguments, the Sequence Recovery Capability of the shuffled frames of the coronary angiography could be an indicator of coupling extent between time and space of antegrade contrast material and hence the TIMI flow grade. Consequently, we proposed that the TSCT in coronary angiography can be used as an indicator of TIMI flow grade. We further proposed the implementation approach to measure the TSCT: to what extent humans can recover the shuffled frame sequence (Sequence Recovery Capability) of coronary angiography.

### 2.4 Ischemia Scores: Stenosis, Sequence Recovery Capability and Their Combination

We featured anatomic structure using stenosis percentage and Functional Capability using Sequence Recovery Capability as illustrated in the above subsection. Finally, we use the combination of these two indicators to be used as a combined indicator indicating Myocardial Ischemia. The combination is in the form:

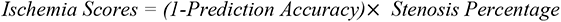

We demonstrated that via combining both anatomic and functional features together to assist the physician to the diagnosis will be superior to using only anatomic features.

### 2.5 The Outcome Event: PCI Conduction after CAG

According to our previous argument, anatomic stenosis doesn’t necessarily cause functional ischemia. However, myocardial ischemia is the root cause of coronary artery disease and its complications which is regarded unobservable.^1^ It is diagnosed through various observable pathological artery structures, symptoms such as stenosis, chest pain etc. However, the challenge is that none of them is demonstrated to be a necessary condition of myocardial ischemia and thus not suitable to act as an indicator of Ischemia. In other words, we so far are not able to judge if myocardial ischemia exists by these observable indicators from the dataset. We thus assumed an Ex Post indicator as the proxy for myocardial ischemia: PCI conduction after CAG. If the patient is conducted the PCI after CAG to relieve the suspicious lesion according to physician’s final decision, we thought he is with the problem of myocardial ischemia. While it exist cases when the physicians make the wrong judgment that the necessary condition is violated and the proxy is thus invalid, we assumed such violation as the outlier in the dataset and they were thus omitted, we developed the further details in the discussions.

### 2.6 An Experiment of the Equivalence between Sequence Recovery Capability and Myocardial Ischemia

We implemented an experiment to validate that temporal and spatial coupling tightness (Recovery Capability) can be an indicator of Myocardial Ischemia. 7 cardiology physicians and 12 ordinary people participated in this experiment. A questionnaire was designed and sent to each of them, the questionnaire sample is attached in Supplemental Material Section 5. They were asked to recover 9 time-shuffled coronary angiographies. The 9 coronary angiographies are distributed as Figure 2. They could be classified into two groups consisting with the outcome event: PCI conduction after CAG and Non-PCI conduction after CAG. However, the group type could not be identified by the subjects during the experiment. Coronary angiography with previous cardiac surgery, previous percutaneous coronary intervention and scheduled PCI were included in the questionnaire, we assumed if the method works on such special and tough cases, it will work on normal cases as well, which strengthen the reliability and adaptivenss of the novel method. Besides, such adaptiveness of the method can simplify the training process of the following automation deep learning model since there is no need to screen the data before feeding them into the model. The model can process various situations of coronary angiography, including those extreme cases that typically excluded out of the dataset. ^6^ We asked the 19 subjects to complete their answers of sequence recovery in 10 minutes independently. We used Cohen’s Kappa^14^ to measure the agreement between the true sequence and the subject’s prediction, the detailed results are illustrated in Section 3.1.

**Figure 2.**
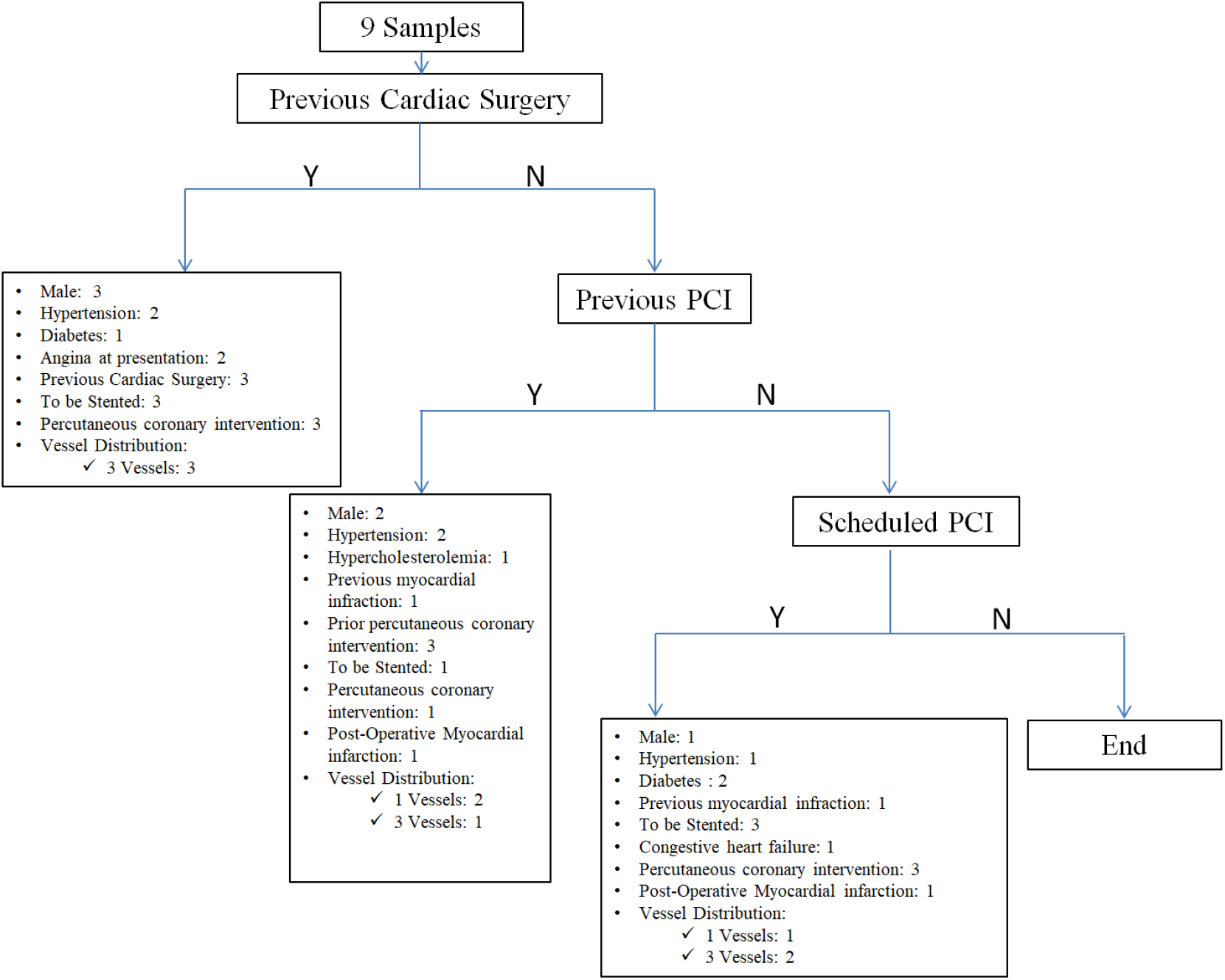
Distributions of 9 Videos Used in the Experiment. There are 9 samples in the questionnaire. 3 videos are from patients with previous cardiac surgery. 3 videos are from patients with previous PCI and 3 videos are from patients with scheduled PCI. These types of extreme cases are used and not excluded in our training set to demonstrate the superior adaptiveness of our method to QFR. As a byproduct, data preprocessing is largely simplified and there is no need to screen the data before feeding them into the automation model which enables the large raw medical data applicable efficiently.

### 2.7 An Unsupervised Deep Learning Model to Recover Sequence Automatically without Labeling

Based on the novel method and its effectiveness, we designed and implemented an unsupervised deep learning model to learn the capability to automatically recover the shuffled sequence of coronary angiography so that the novel method is able to be automated and scaled. By a self-supervised approach to shuffle a certain proportion of the frame sequence and generate labels automatically depending on whether the current frame is disordered, the deep learning model works without any labeling efforts. Besides, it is not necessary to screen and clean the input training set since the coupling tightness theory is proved to be correct even for some interfered cases such as coronary angiography with previous cardiac surgery, previous percutaneous coronary intervention (PCI) and scheduled PCI. The feature enables the large raw coronary angiography data applicable without heavy labeling.

Deep learning has achieved great success in computer vision, particularly in the field of medical. It is well known that the first application of deep learning is medical image processing, including magnetic resonance images of the brain, ultrasound images of breast nodules, CT and X-ray images of the lungs, which reflect the structure of organs and the attributes of lesions through continuous groups of images from different angles.^15^ However, deep learning in coronary angiography is very limited, mainly for the two reasons as follows:

1. CAG is exhibited in a form of videos. There are several studies of deep learning based on coronary angiography, in which only the certain frames from the coronary angiography video are selected in the further analysis. The static assessment of the lesion from the certain frames can only get anatomical features. After all, anatomic stenosis is not equivalent to the true ischemia. Such property restrains the influence and reliability of such studies.^16^
2. The annotation of medical images consumes huge medical resources, if such annotation can only be completed by professional doctors, in particular. Since coronary angiography is exhibited in a form of videos, with several frames, the annotation will thus consume even more resources, causing the previous studies, even based on coronary angiography, to select key frames from the whole video to annotate and analysis. Such static methods lose the tremendous information advantage of coronary angiography. Other related studies just use CCTA instead of CAG and are constrained to the very small sample size probably due to the huge consumption of labeling engineering. Our proposed model is totally label-free. Therefore it is very easy to be implemented and generalized in real life.

We designed and implemented an deep learning model to learn the capability of sequence recovery, we highlighted that owing to the discovery that sequence recovery capability is an indicator of TIMI flow grade, the deep learning model is based on dynamic changes of the antegrade contrast agent of coronary artery, resulting from a functional view of stenosis severity detection, besides, it is completely unsupervised and no labeling effort is needed, enabling the clinical application relying on huge medical data.

#### 2.7.1 Training Set

From January 1st 2016 to December 31th 2016, we collected 1980 unidentified videos of Right Coronary Artery (RCA) coronary angiographies. Notice that we included some extreme cases that typically excluded from the dataset to complete the whole analysis as we assumed them not violating the working conditions of the model. For example, patients with previous cardiac surgery, previous PCI and scheduled PCI are not excluded from the training set since the antegrade contrast agent dynamics are the same as typical coronary angiography. And we thus proved the strong adaptiveness of DZL to the vessels tree restructure cases where QFR is dragged. We use 1312 videos as the training set to train the model.

#### 2.7.2 Test Set

We collected additional 668 videos of RCA coronary angiographies as the test set. The main performance results are given in Section 3.2.

#### 2.7.3 The Model

We first shuffled the video sequence of coronary angiography to generate the self-supervised labels automatically, then we generated an effective optical flow zone to calculate on. Lastly, an Encoder-Decoder GRU neural network^17, 18^ is used to learn the capability to recover the sequence. The model prediction accuracy was used as a proxy to the Recovery Capability and hence an indicator of Myocardial Ischemia. The lower prediction accuracy means weaker Sequence Recovery Capability. The details of the deep learning model are given in Supplemental Material Section 2.

### 2.8 Use DZL to Get Functional Evaluations of Vessel Segments: Difference DZL

So far, we have argued the prediction accuracy of model DZL could be used as a proxy for Myocardial Ischemia to functionally evaluate the coronary artery. We then exerted the effort to measure the functional capability of local segments of coronary artery. The successful tackling to measure the vessel segments functional capability locally is crucial for real-time procedure assistance. Physicians are illustrated the Hemodynamics Ratio of any part of the coronary artery and such information is valuable to help them make the final diagnosis and treatment decisions.

For the suspicious coronary artery segments, the insight is to compulsorily occlude the segments by setting the pixels from the area to 0 for each frame of the coronary angiography video. Such operations act like artificially occluding the suspicious part and it increases the difficulty to recover the certain shuffled frames when the antegrade contrast agent passes through the factitious occluded area and thus increase the difficulty to recover the overall video. Two cases will consequently happen. If the suspicious coronary artery segment is functionally fine, there will be a larger gap of Difference DZL scores between occluded or not. If the suspicious coronary artery segment is functionally bad, the corresponding gap will be much smaller and ideally near zero if the suspicious one is originally occluded.

## 3 Results

### 3.1 The experiment

The Cohen Kappa matrix between subjects’ predictions and the true sequence are listed in Table 1. From the results, the agreement between predictions and the true sequence is higher in PCI conduction group than Non PCI conduction group no matter the subject is physician or ordinary people, besides, physicians’ prediction agreements are overall higher than ordinary people in both groups as physicians have affluent clinical experience and practice to professionally judge the coronary angiography status. The detailed confusion matrixes of physicians and ordinary people are given in Supplemental Material Section 4. The overall AUC achieved in the experiment from 7 professional physicians is 0.92.

**Table 1.** Cohen Kappa Matrix between Subjects’ Predictions and True Sequence. The Cohen Kappa of Non PCI conduction group is overall higher than PCI conduction group. The Cohen Kappa of physicians group is overall higher than ordinary group. In other words, severe lesions will cause worse sequence recovery consistency and physicians are more skilled than ordinary people to distinguish the shuffled order. Besides, the AUC of physicians is up to 0.92, validating a competitive performance and potential challenging the QFR approach.

|  | **Physicians** | **Ordinary People** |
| --- | --- | --- |
| **PCI conduction Cohen Kappa** | 0.48 | 0.25 |
| **Non PCI conduction Cohen Kappa** | 0.89 | 0.56 |
|  | **Physicians** |  |
| **AUC** | 0.92 |  |

### 3.2 Automation Model

For the automation approach of the DZL method, the results of the model using the 1312 videos training set and the 668 videos test set are listed in Table 2. We validated the truth that comprehensive consideration over both anatomic and functional perspectives is worth better clinical results for the diagnosis of myocardial ischemia. For Difference DZL, figure 4 shows the operations and effectiveness of Difference DZL.

**Table 2.** Main Performance Results.

|  | **Ischemia**<br>**Scores** |
| --- | --- |
| **AUC-median--%**<br>**(95% CI)** | 84<br>(80.69-87.02) |
| **Sensitivity**<br>**-- %(95%CI)** | 80<br>(75.84-83.77) |
| **Specificity**<br>**-- %(95%CI)** | 86.92<br>(82.06-91.37) |
| **Positive**<br>**predictive value**<br>**-- %(95%CI)** | 90.45<br>(86.43-94.02) |
| **Negative**<br>**predictive value**<br>**-- %(95%CI)** | 73.66<br>(68.17-78.73) |

## 4 Discussions

### 4.1 Non-Invasive, Real-Time. Adaptive Evaluation for Functional Ischemia

In terms of the coronary artery disease causing by myocardial ischemia, diagnosis from anatomy branch facing the challenge from functional branch as solid clinical evidences are demonstrated that anatomy stenosis does not necessarily mean functional myocardial ischemia.^2^ However, although functional measurements such as FFR is attained in real-time procedure, it is invasive and will bring more operation time and risks. Besides, the non-invasive QFR is also attained in real-time procedure, it is not easy to adapt to post cardiac surgery or operation scenario as both cardiac surgery and operation will restructure the vessels tree and thus change the hemodynamics and violet the validness of the predictions from QFR. (Figure 3) As far as we know, our method is the first to assist real-time revascularization procedure to make the treatment decisions as well as prompt prognosis evaluation after the revascularization in a non-invasive manner and applicable to patients with cardiac surgery or operation history. The whole calculations are within 1 minute.

### 4.2 Dynamic Based on Coronary Angiography

In terms of diagnostics based on coronary angiography, as far as we know, all the existing researches are static based on anatomy. As a typical example, Moon et al. (2021)^19^ used CNN to achieve the automatic recognition of coronary lesion based on coronary angiography. However, the preprocessing technique of their method was to select key frames instead of utilizing the whole coronary angiography video. Besides, their method was a purely supervised learning which needs manual label efforts a lot. Analysis based on static image corresponds to anatomy diagnosis while a stenosis found in a static image doesn’t necessarily mean ischemia.^16, 23^ Our model is based on analysis of the whole coronary angiography video calculating in a TIMI flow grade perspective instead of separated images which is theoretically dynamic in a functionally perspective.

**Figure 3.**
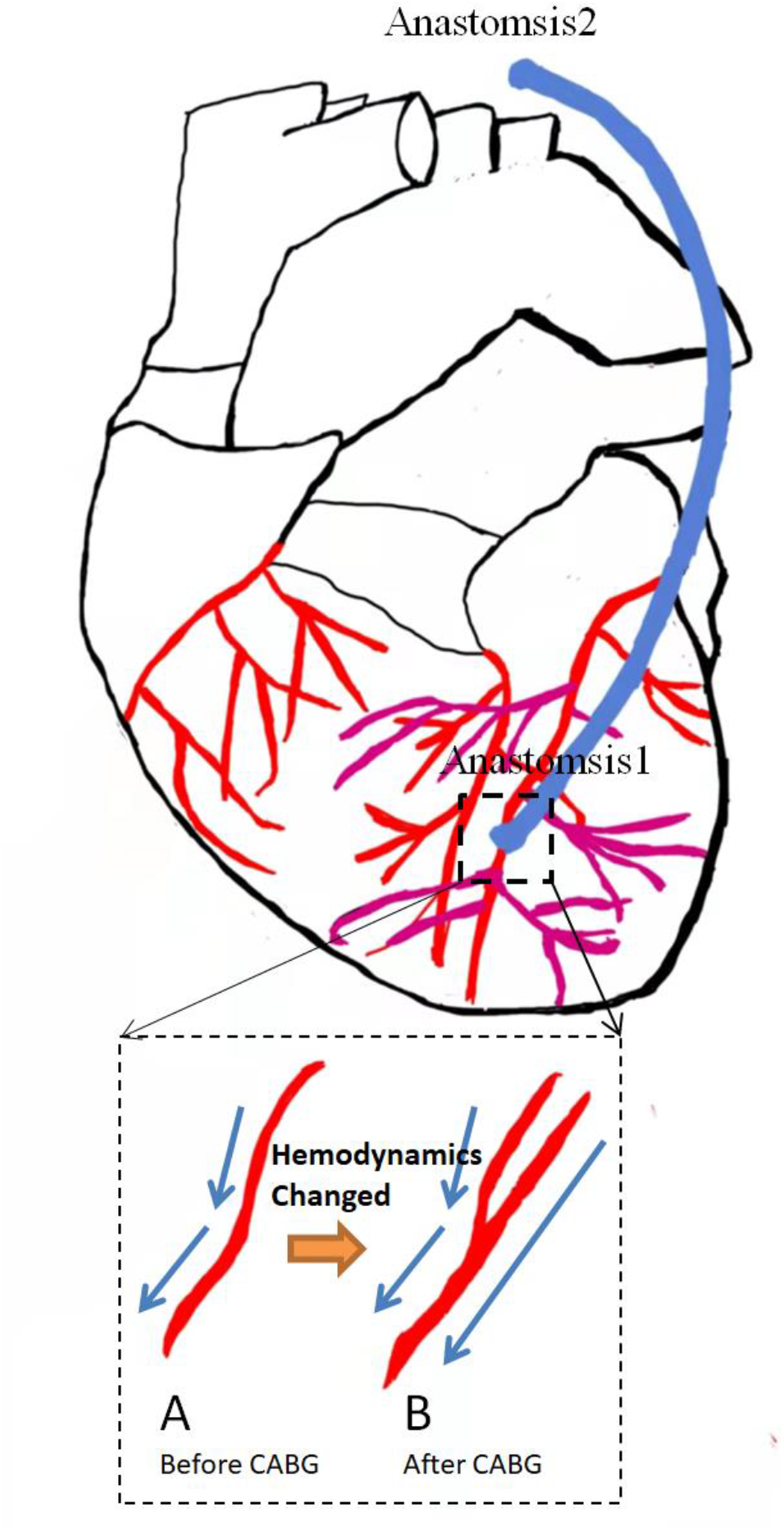
Hemodynamics Change Before and After CABG. The anastomosis1 was made during a CABG. It extended a branch to the original vessels tree and thus changed the whole hemodynamics. Anastomosis2 was not illustrated as it follows the same logic. The change of hemodynamics will weaken the effectiveness of prediction models like QFR which to some extent relies on prior vessels tree information.

**Figure 4.**
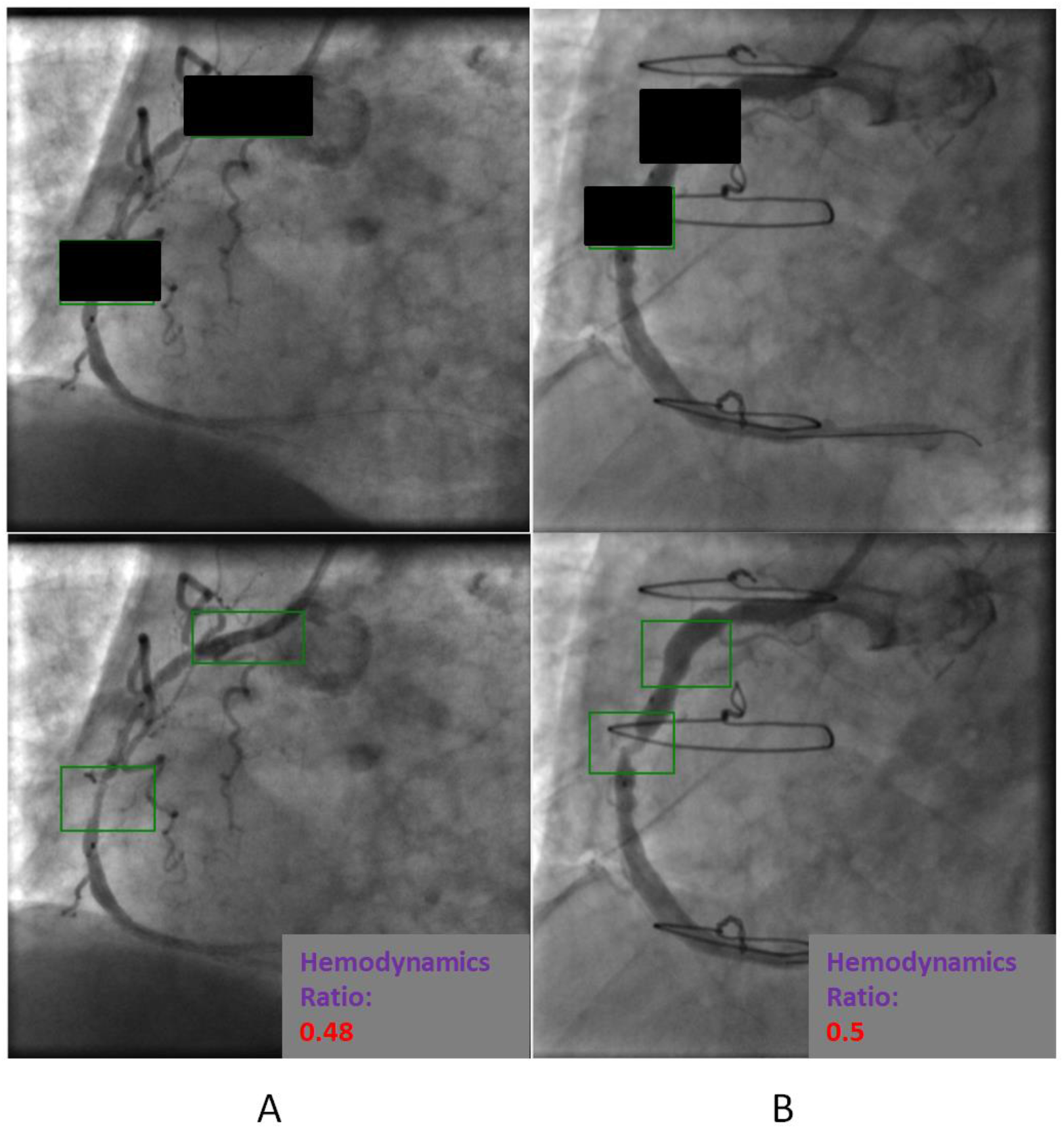
Difference DZL. There are two green frames. The upper one outlines the healthy proximal vessel segment and the lower one outlines the lesion in middle vessel segment. The Hemodynamics Ratio shows the functional capability value of the lesion segment over healthy segment. A is the sub-figure for a patient to be conducted PCI and B is the sub-figure for a patient with CABG history, the two cases are typical to test the adaptiveness of DZL where QFR is dragged. By setting the pixel values in the interested frames to 0 along the whole video, the difference DZL achieves the hemodynamics ratio of 0.48 and 0.51 for A and B respectively, indicating a near half functional capability for the lesion segment comparing to healthy segment.

### 4.3 Label Free

We automated our novel method using an unsupervised learning model which is another breakthrough and contribution to related literatures and applications. There are many mature annotation libraries in image recognition, the most famous one is ImageNet^24^, and there are some other annotation libraries in video recognition such as Kinetics. These databases cover general scenarios. However, in the field of medical imaging, especially coronary angiography, there is no mature annotation library developed. For studies using deep learning techniques in this field, researchers need to complete the label engineering work by themselves which is highly costly since the annotation work must be finished by professional doctors.^16, 19,20,21,22^ This makes the development of deep learning, especially supervised learning in medical image field greatly limited. According to statistics by May 2020 from OECD (Organization for Economic Cooperation and Development), the average annual income of a specialist in hospitals of United States is 350300 dollars, and for a GP is 242400 dollars. That is to say, a specialist earns up to $168 an hour and a GP earns up to $116 an hour, even without taking into account national holidays. Doctors estimate that it takes about 15 to 20 minutes to fully evaluate a coronary angiography, so even if a more junior GP to do the annotation, the cost is as high as $30 for each coronary angiography. For deep learning, a lot of labeled angiographies are needed. For example, in this paper, 1312 videos of RCA angiography were used for training and 668 for testing, and the cost of manual labeling alone was at least as high as $44080, not to mention some larger training program. In our study, there is no need to manual labeling at all so that the training cost is greatly saved and large-scale training becomes possible. (Supplemental Figure 6)

### 4.4 Potential of the DZL Automation Model

From the performance of final results, we can see that the automation model still has great potential to improve. The automation model used optical flow technology to represent sequence feature between frames which is an idea of dimension reduction. The dimension of the model input reduces by 2621.44 times which is very considerable while the performance of the model is maintained in a good level. However, dimension reduction will eventually bring about the loss of information and indirectly affect the final AUC. If we can enhance the parallel computing power of computers, such as using GPU instead of CPU to design and implement the model so as not to reduce the dimension too much, it will thus improve the final AUC.

### 4.5 Advance the Development of Retrospective Medical Study

Since we provided a novel method to functionally evaluate coronary artery based on coronary angiography video, which means such huge amount of history data could be utilized to conduct functional re-evaluation and attain updated diagnosis. If the previous diagnosis and updated diagnosis are not consistent, it is worthwhile to take a deeper investigation and figure out the reasons and outcomes behind. The possibility released by our method to functionally re-evaluate the huge history coronary angiography videos without angle constraint in QFR will heavily prosper the retrospective medical study based on various counterfactual evaluations and push the development of clinical practice forward to a great extent.

### 4.6 Limitations

In this study, we used an Ex Post indicator (PCI conduction after CAG) as the proxy for myocardial ischemia. However, there could be some misdiagnosis from physicians. Although our data came from a leading hospital in Shanghai, China, such misdiagnosis was hardly to happen, the outliers were possible to appear still and bias our results to some extent. Further work could use samples with FFR and QFR data to exactly measure the myocardial ischemia extent. Next, further FFR/QFR data is also needed to validate our Difference DZL since the actual ischemia status of vessel segments is necessary to be measured and set as a baseline. Also, only the Right Coronary Artery (RCA) Coronary Angiography data was used in the experiment and DZL automation model. However, the method is general and not constrained to any coronary artery parts. It can also be applied to Left Coronary Artery (LCA). Future work can further validate, improve and extend the DZL experiment and automation model. Lastly, in this study, we skipped all the traditional data preprocessing procedures in order to demonstrate the pure power of DZL and its automation. However, it also constrains the performance of DZL. Future work can gradually add the typical preprocessing procedures such as segmentations, centerline extractions etc.

## 5 Conclusion

As an old Chinese saying: “A swift horse is common, but a bole is not”. The “Sequence Value” of coronary angiography is buried deep. It is just like a swift horse that can lead a revolution in the realm of coronary artery disease diagnosis and treatments, it even gains the potential to overshadow QFR to functionally evaluate coronary artery in a non-invasive, read-time, adaptive manner. Since QFR was successfully applied in practice and demonstrated huge clinical value. Recent research has showed its advantage over using CAG to guide the revascularization procedures that via using QFR instead can reduce 35% post-operation risk.^1^ Our study aimed to challenge the champion as we recognize the “Swift Horse” and its tremendous medical value. We thus proposed DZL as well as its automation. In our experiment, the AUC of DZL reaches 0.92 which is not quite inferior to QFR. Although its automation model reduces the performance to 0.84, it is totally unsupervised and no labeling effort is needed which is a breakthrough in the field since the unsupervised property means huge medical labor saving and large clinical application feasible. We further proposed Difference DZL to measure the functional capability of specific coronary artery segments to guide the revascularization procedures. Both DZL and Difference DZL work in a real-time fashion and can be conducted within 1 minute. In a nutshell, the “Swift Horse” is leading a revolution in functional evaluation of coronary artery disease and we would like to invite boles all over world to dig the gold mine.

## Abbreviations

QFR: Quantitative Flow Ratio
FFR: Fractional Flow Reserve
TSCT: Temporal and Spatial Coupling Tightness
TIMI: Thrombolysis In Myocardial Infarction
RCA: Right Coronary Artery
LCA: Left Coronary Artery
DZL: Disarranged Zone Learning
LV: Left Ventricle
CMR: Cardiac Magnetic Resonance
CT: Computed Tomography
CCTA: Coronary Computed Tomography Angiography
CAG: Coronary Angiography
PCI: Percutaneous Coronary Intervention
CABG: Coronary Artery Bypass Grafting
RNN: Recurrent Neural Network
GRU: Gated Recurrent Unit
ROC: Receiver Operating Characteristic
AUC: Area Under the Curve
GPU: Graphic Processing Unit

## Data Availability

All data produced in the present study are available upon reasonable request to the authors

## 1. Paper Definitions

### 1.1 Definition of Optical Flow

Optical flow is the instantaneous motion speed of the pixels of a spatially moving object in the observed imaging plan. Optical flow uses the change of pixels in the image sequence in the time domain between adjacent frames to calculate the motion information of objects between adjacent frames. Optical flow can be generally divided into two categories: sparse optical flow and dense optical flow.

### 1.2 Definition of machine learning and deep learning

Machine Learning uses mathematical operation to mimic human’s capability to make specific judgments, typically the classification decisions. It uses huge amounts of data samples as input, through the statistical model and output the classification labels. Deep learning is a subset of machine learning, using multi-layer neural network to make the predictions. There are two schemas of machine learning, supervised learning and unsupervised learning.

### 1.3 Definition of supervised learning

The training process is guided by true labels as feedback to adjust the model parameters, such schema is termed as supervised learning. In medical realm, supervised learning consumes huge labeling cost as the labeling can only be judged by professional physicians. Especially in assisting medical image analysis, the labeling efforts are heavy as human takes time to digest visual high dimensional information.

### 1.4 Definition of unsupervised learning

The training process is conducted without relying on true labels. The model parameters are adjusted through figuring out the intrinsic representation of data itself by various mathematical operations, such as clustering the data samples that are similar to each other. Such schema is termed as unsupervised learning. We proposed an unsupervised learning for media of medical video by shuffling the video frame sequence and generating the self-labeling automatically. The method intrinsically takes use of the sequential features covered in the video to cluster the video frames into two categories of shuffled or not shuffled. The whole process is not relied on any manual labeling work and can be applied to huge data scale.

### 1.5 Definition of Recurrent Neural Network

A Recurrent Neural Network (RNN) is specified to be used to process sequential data. Instead of defining parameters at each time step, the RNN uses a parameter sharing framework to keep fixed length of parameters matrix updated along the sequence so that the RNN can generalize the sequential feature concealed in the sequence and make corresponding predictions. For example, given the context word sequence and predict the next word. Basically, the RNN typically handles the classification problem as well except the classes to be distinguished are extended to hundreds or thousands. In this paper, we applied the RNN to process the image sequence, especially the optical flow sequence of the coronary angiography.

### 1.6 Definition of Encoder Decoder Sequential Model

The Encoder and Decoder Sequential Model is first proposed by Sutskever [1] to handle the variable length sequential input and predict an un-fixed length sequential output. It is a typical model to handle sequential inputs. The core RNN inside model can be LSTM, GRU, etc. Since the length of the optical flow of each frame image is not fixed depending on the sequence length of each coronary angiography, we chose this model to apply in our scenario.

### 1.7 Definition of GRU Model

It is a kind of Recurrent Neural Network inspired by the LSTM [2], but it is much simpler to be designed and implemented, it has comparable or even superior performance than LSTM in various applications.

GRU has two gates: reset gate *r_j_* and update gate *z_j_*, the activation of j-th hidden unit is computed as:

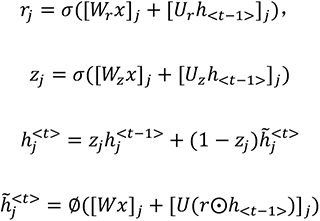

σ is the sigmoid activation function, and [.]*_j_* denotes the j-th element of a vector. x and *h_t_*_-1_ are the input and the previous hidden state, respectively. *W_r_* and *U_r_* are weight matrices which are learned.

### 1.8 Definition of attention mechanism

The attention mechanism can be regarded as an additional layer of the neural network to multiple the attention weight from. It equals to weight the hidden unit of each time step to construct the fixed-length vector according to current decoder hidden output. Those with higher similarity scores of each source hidden state and current decoder hidden state will be attained higher weights and vise versa. Consequently, the fixed-length vector turns to be a dynamic update of the combination of attention weights and source hidden states. We used the global attention approach that the fixed-length vectors are calculated from combinations of all source hidden states and their weights. The intuition behind our attention mechanism is that those optical flow coordinates will show disparate pattern between shuffled and un-shuffled frames. When the model classifies the output, it should take more use of the information generated by its own class and slightly omit the opposite class samples.

### 1.9 Definition of clip gradient

Clip gradient is an approach to restrain the gradient within a certain maximum to prevent the model backpropagation process from exploding gradient event. Our clip parameter is set to 50 in our model.

### 1.10 Definition of Bootstrapping Procedure

Bootstrapping was used only to estimate 95% confidence intervals (CI) for the performance metrics of our classification results (i.e., AUC, sensitivity, specificity). We applied n-out of-n bootstrap with replacement from our dataset. For each bootstrap sample, we calculated and reserved the performance metrics for that bootstrap sample. The bootstrap sampling was repeated for 2000 times. We then estimated the 95% CI by using the 2.5 and 97.5 percentiles of the empirical distribution of corresponding metrics. [3]

### 1.11 Definition of Accuracy and predictive values calculations

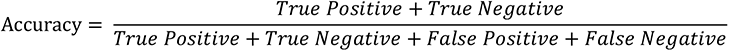

True positive: The prediction is correct and the actual value is positive.

False positive: The prediction is wrong and the actual value is negative

True negative: The prediction is correct and the actual value is negative

False negative: The prediction is wrong and the actual value is positive

Sensitivity: Sensitivity is the proportion of true positives tests out of all patients with a condition. [4] In other words, it is the ability of a test or instrument to yield a positive result for a subject that has that disease. [5]

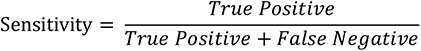

Specificity is the percentage of true negatives out of all subjects who do not have a disease or condition [4]. In other words, it is the ability of the test or instrument to obtain normal range or negative results for a person who does not have a disease. [5]

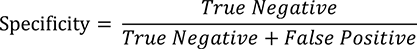

PPVs determine, out of all of the positive findings, how many are true positives; NPVs determine, out of all of the negative findings, how many are true negatives. As the value increases toward 1, it approaches a ‘gold standard. [6]

The formulas for PPV and NPV are below.

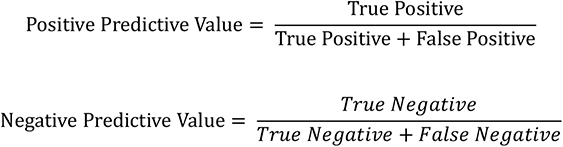

## 2. Development of the Encoder Decoder GRU Sequential Model

### 2.1 Step 1: Disarranging Sequence Order of Coronary Angiography

A coronary angiography video is composed of many frames and each frame is an image which records development state of the vessels at a certain time when the contrast agent is injected. We first shuffled the frames sequence of the video. We set θ, the Disarranged Ratio, equal to 0.5. In other words, half of the frames in a video is shuffled while the other half is not shuffled. If the total number of frames in the video is odd, we round the half to be shuffled.

### 2.2 Step 2: Zone Generating

We were concerned about the dynamic process of blood vessels during contrast injection. We used optical flow technology to extract sequence features and formed an effective zone to calculate on.

Optical flow is the instantaneous motion speed of the pixels of a spatially moving object in the observed imaging plan. Optical flow uses the change of pixels in the image sequence in the time domain between adjacent frames to calculate the motion information of objects between adjacent frames. Optical flow can be generally divided into two categories: sparse optical flow and dense optical flow. Lucas and Kanade(1981) [7] is a typical method to calculate the sparse optical flow. This method needs to use some method to locate the corner points as the initial points of the optical flow. We used Shi and Tomashi(1994) [8] method to locate the corner points but find these points are less related to vessels due to various disturbances such as spinal pixels, heartbeat, angle transformation etc. That was, the corner points were not generated on the vessel, so the generated optical flow failed to represent the dynamic effect of the contrast agent injection process. So we chose to compute the dense optical flow proposed by Farneback(2003) [9]. It computed the optical flow of all pixels between frames. It was mainly through polynomial expansion and the assumption of pixel displacement invariance to reach the optimal optical flow.

We then encountered the problem of further selection of effective feature points and formed an active zone to enforce the disarranging function on. Because dense optical flow calculated the optical flow of all pixels in a frame, only a small part of which was related to blood vessels. If all the pixels were analyzed: (1). A compensation mechanism would work which put a negative impact on the analysis of the results. Since coronary angiography also recorded the movement of part of the spine, heartbeat as well as some movement of the camera angles, so it might make it easier to recover the disorder via using such additional information though the vascular visualization part was not easy to be recovered. Bias might correspondingly be generated. (2). the dimension for Farneback optical flow of each video was up to 262144, which would encounter the curse of dimensionality.

It thus was necessary to develop a screening mechanism to select an “effective zone” from where the optical flows were generated from the pixels of vessel.

Supplemental Figure 1 was a dense optical flow calculation process, from which we got such a view that the points associated with blood vessels generally changed sharply in color and brightness. From the visualization process, different color and brightness expressed different magnitudes and angles of optical flow.

Supplemental Figure 2 was a vessel in polar coordinate plane. We finally selected the points whose flow magnitude variances were above 80 percentile and used them to form an effective zone. Moreover, we randomly selected 100 points from the 52428 effective points as the final zone. The final zones and its optical flow traces were illustrated in Supplemental Figure 3, column C and column D. All the hyperparameters are decided based on trial and error on validation set.

### 2.3 Step 3: Learning

The objective of the learning model is to learn the capability to recover disordered frames like human. To simplify the model, we designed the model as a two-class classification model, that is, we trained the model to tell if the specific frame is disarranged or not. The labels guiding the training process were generated automatically comparing to the original order. Once the model was trained to learn the recovery capability, its accuracy was used as an approximator to Recovery Capability (TSCT) and therefore be an indicator of TIMI flow grade and myocardial ischema. The model training process of learning recovery capability worked in a self-supervised approach. In other words, it is not necessary to know the “answer” of TIMI flow grade beforehand, the model intrinsically worked in an unsupervised learning approach.

In terms of the learning of recovery capability, as our input was the sequence of disarranged optical flow from the final effective zone, we chose the recurrent neural network (RNN) which was specifically used to learn and predict the sequence. Sutskever et al.(2014) [1] found that using two separate RNN can be very good realization of the sequence-to-sequence prediction. In our model, the RNN we chose is GRU (Gate Recurrent Unit) invented by Cho et al.(2014) [2] Such architecture handled variable-length inputs corresponding to variable-frames-counts of different angiography videos. One RNN was for the encoder that received variable-length sequence inputs (max length×batch size) and transferred the inputs into a fix length vector (max length×batch size×hidden size). The input to each GRU module was a sequence element, the output was an output vector and a hidden state (n layers×batch size×hidden size). The other RNN was the decoder, its each GRU module took a sequence element and the fixed length vector from encoder as input and output a prediction of the next sequence element (batch size×class counts) and the hidden state to be passed to its next GRU (n layers×batch size×hidden size). The overall Encoder-Decoder GRU architecture was illustrated as Supplemental Figure 4.

## 3. Experiment Statistical Data

The statistical data of the confusion matrix of 7 physicians and 12 ordinary people is illustrated in Supplemental Table 1, Supplemental Table 2, Supplemental Table 3 and Supplemental Table 4.

## 4. Figures and Tables

**Supplemental Figure 1.**
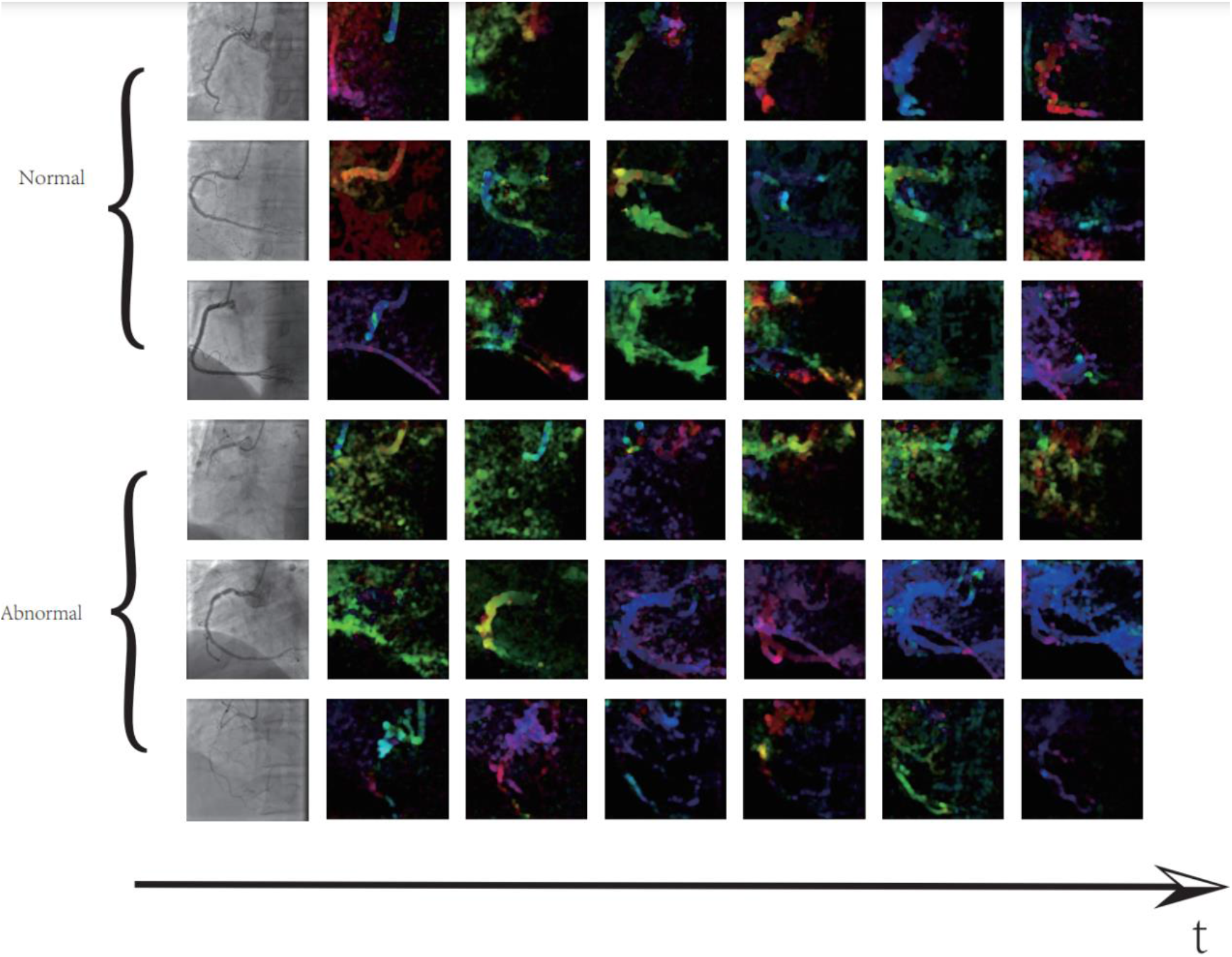
Dense Optical Flow Calculation Process. The first three rows are from 3 normal coronary angiographies, the last three rows are from 3 abnormal coronary angiographies. The black arrow is the time axis. The dense optical flow is developed along the direction of time axis.

**Supplemental Figure 2.**
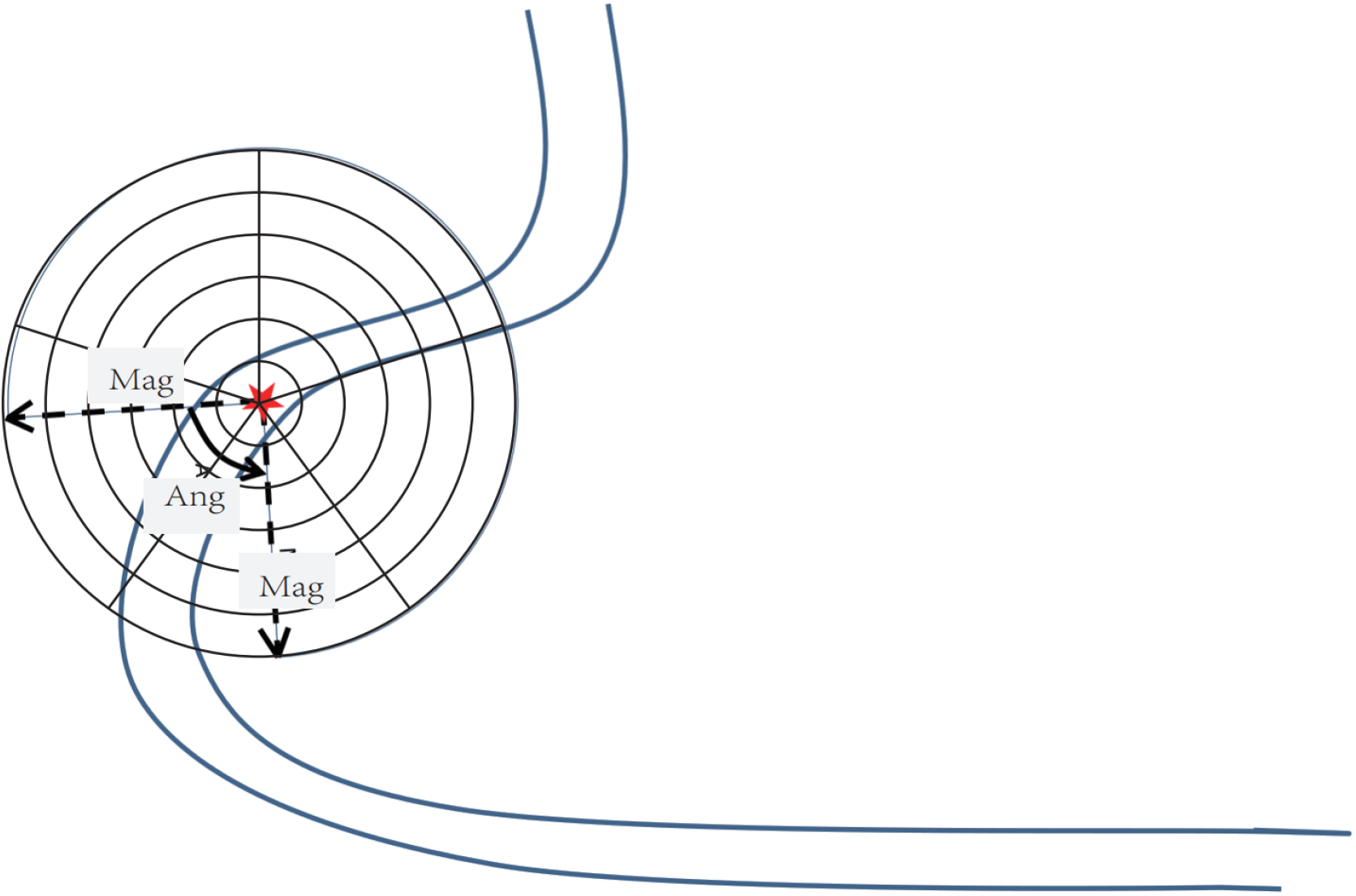
Polar Coordinate Plane View. For the current pixel point (the red star) on the RCA vessel, the dense optimal flow is developed depending on Magnitude (Mag) and Angle (Ang). Different Mag and Ang will cause different color and brightness of the pixel.

**Supplemental Figure 3.**
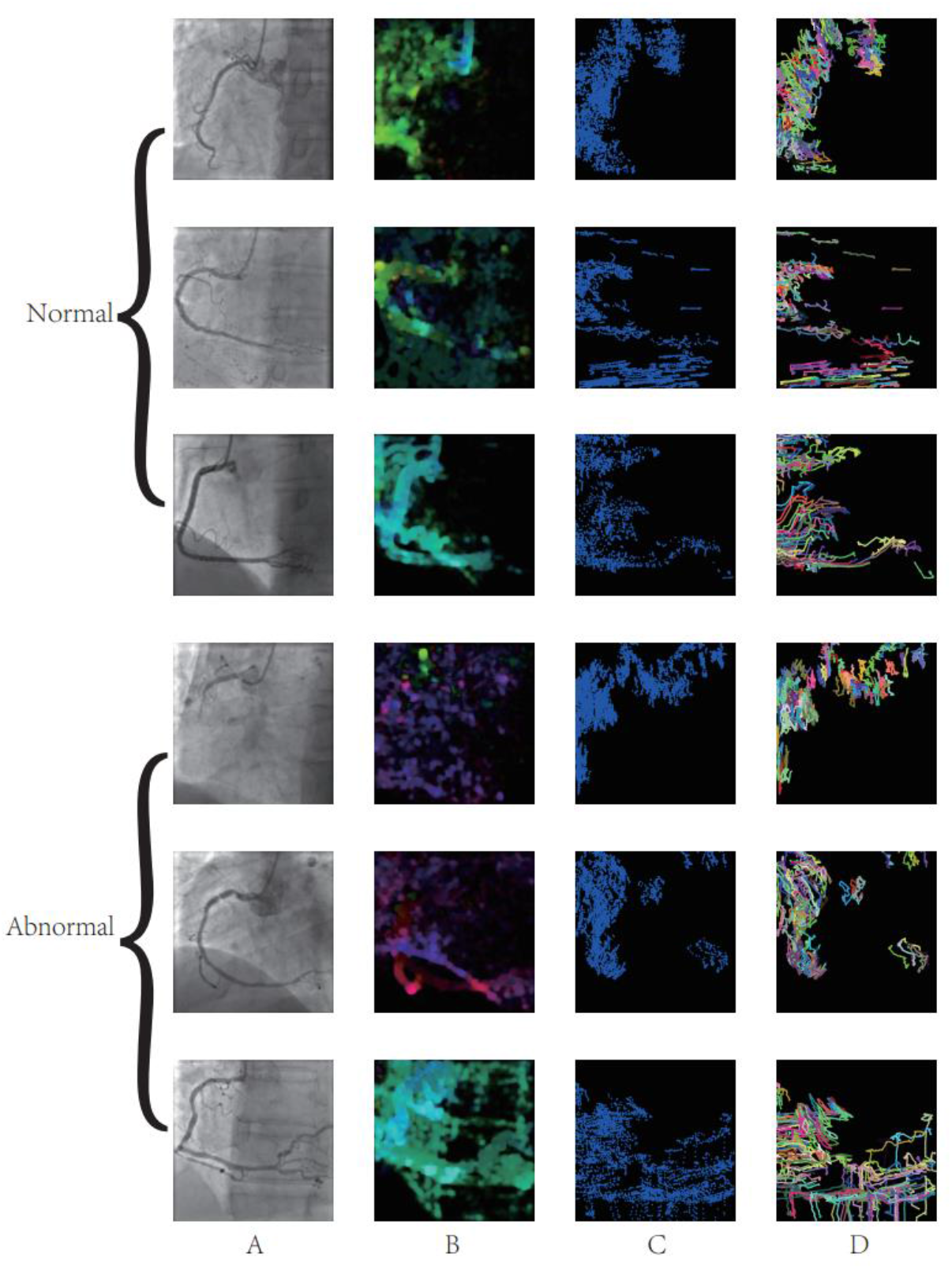
Effective Zone Generation. The first three rows are from 3 normal coronary angiographies, the last three rows are from 3 abnormal coronary angiographies. Column (A) are from the original angiographies. Column (B) are the calculation results of Farneback optical flow of the frames in the segment. Column (C) are the randomly selected 100 optical flow points from the optical flow point set above 80% percentile of magnitude variance. Column (D) are the optical flow traces from 100 points selected.

**Supplemental Figure 4.**
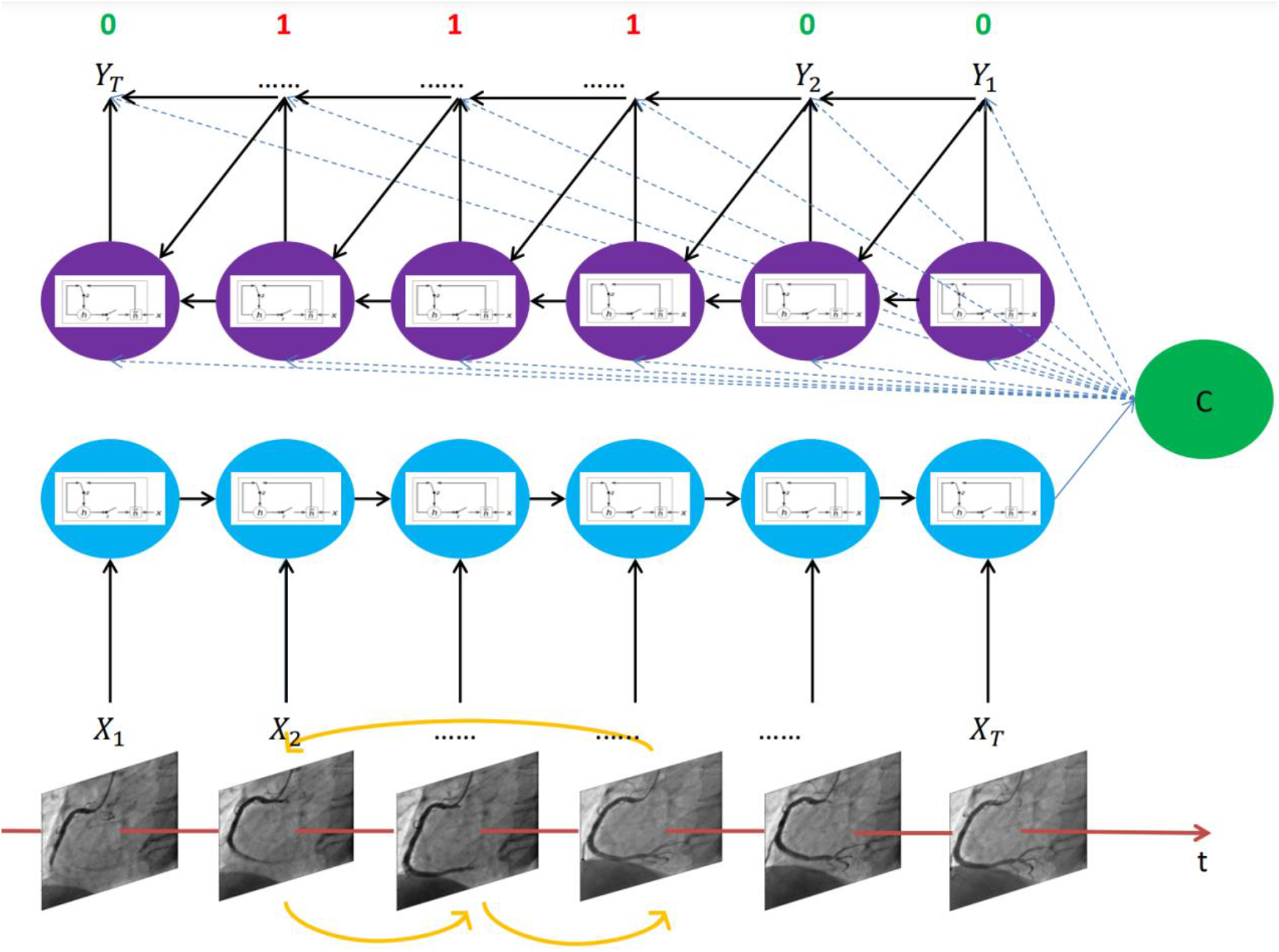
Encoder-Decoder GRU Model. The bottom block is effective zone of disarranged frames as the input of the Encoder-Decoder GRU model. The X block is the encoder module, each node is a GRU unit. The green circle C is the fixed length vector output by the encoder. The y block is the decoder module, each node is also a GRU unit. The loss of the model is calculated through the output of the Y block and the binary labels are generated automatically via the disarranging status of the input frames. If the specific frame is disarranged, it is correspondingly labeled as 1, otherwise, labeled as 0.

**Supplemental Figure 5.**
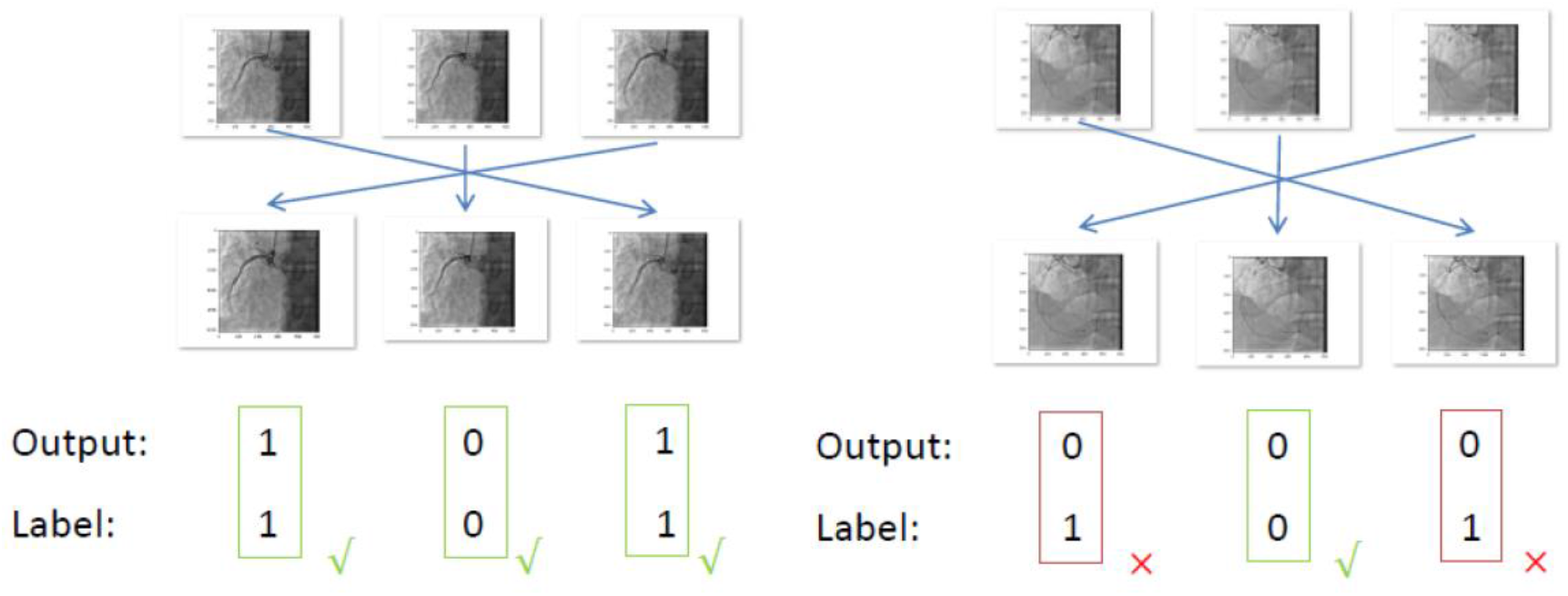
The Intuition. The sub-left figure is the illustration of a sequence of frames from a video of normal right coronary artery. For example, the first and third frames are exchanged and the second frame is not moved. Hence, such disarranging causes the first and third frames labeled as 1 and the second frame labeled as 0. For the sub-right figure, the settings are the same except that it is an abnormal and occluded right coronary artery. We argue the normal coronary angiography is with higher TSCT and high model prediction accuracy. The example shows that the sub-left output-label pairs are consistent, indicating a high model prediction accuracy. Meanwhile, the sub-right output-label pairs are more different, indicating a lower model prediction accuracy.

**Supplemental Figure 6.**
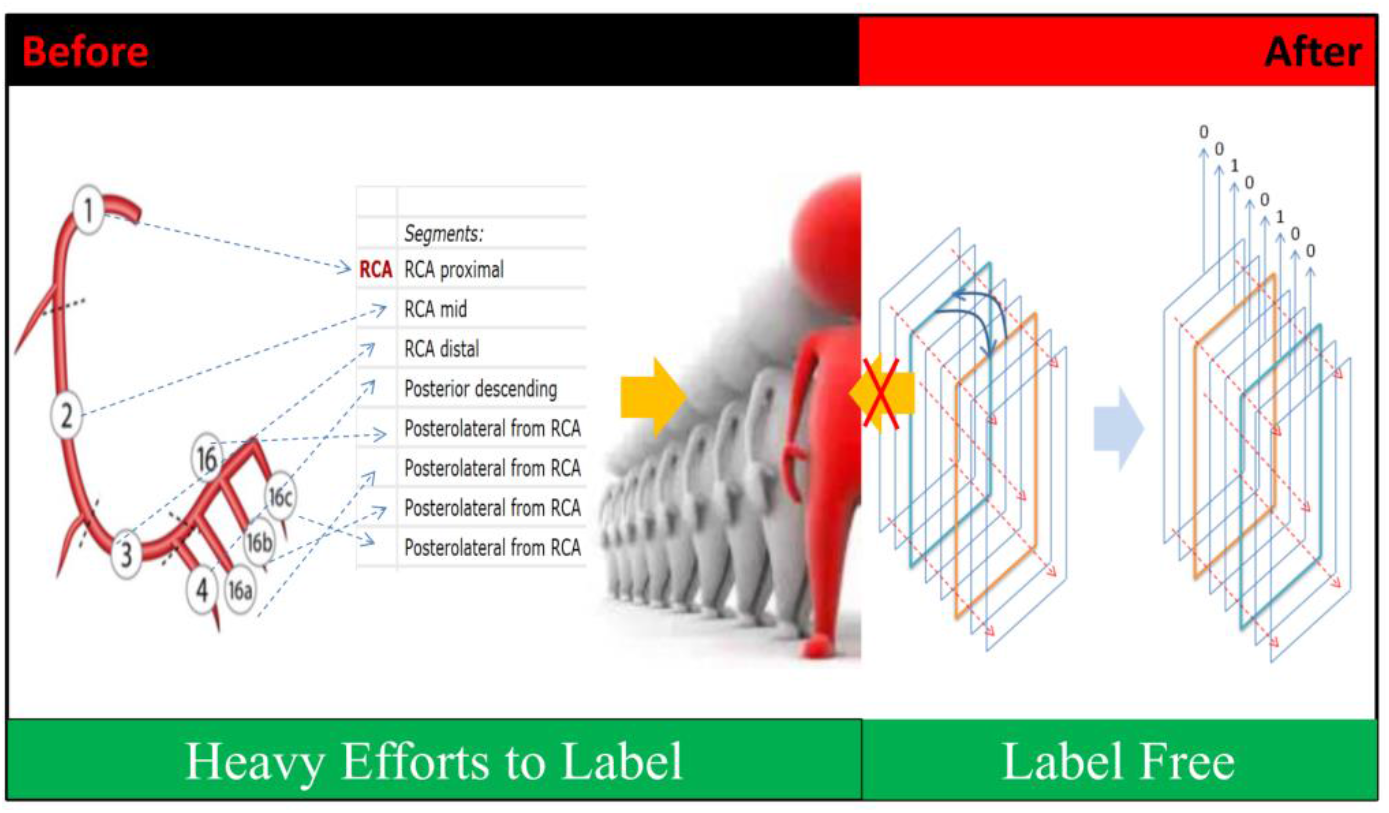
The Laborious Efforts Saved Before and After. From the Syntax Score scoring process, in the right coronary artery (RCA), there are 8 positions to potentially have lesions. Hence there are 256 combination possibilities to be diagnosed as an abnormal vessel. That is to say, to label the RCA, doctors need to exclude the 256 possibilities so as to make a final decision of a normal vessel. Hence the labeling efforts are laborious. On the other hand, the model in this study is 100% label free and can be generalized very easily.

**Supplemental Figure 7.**
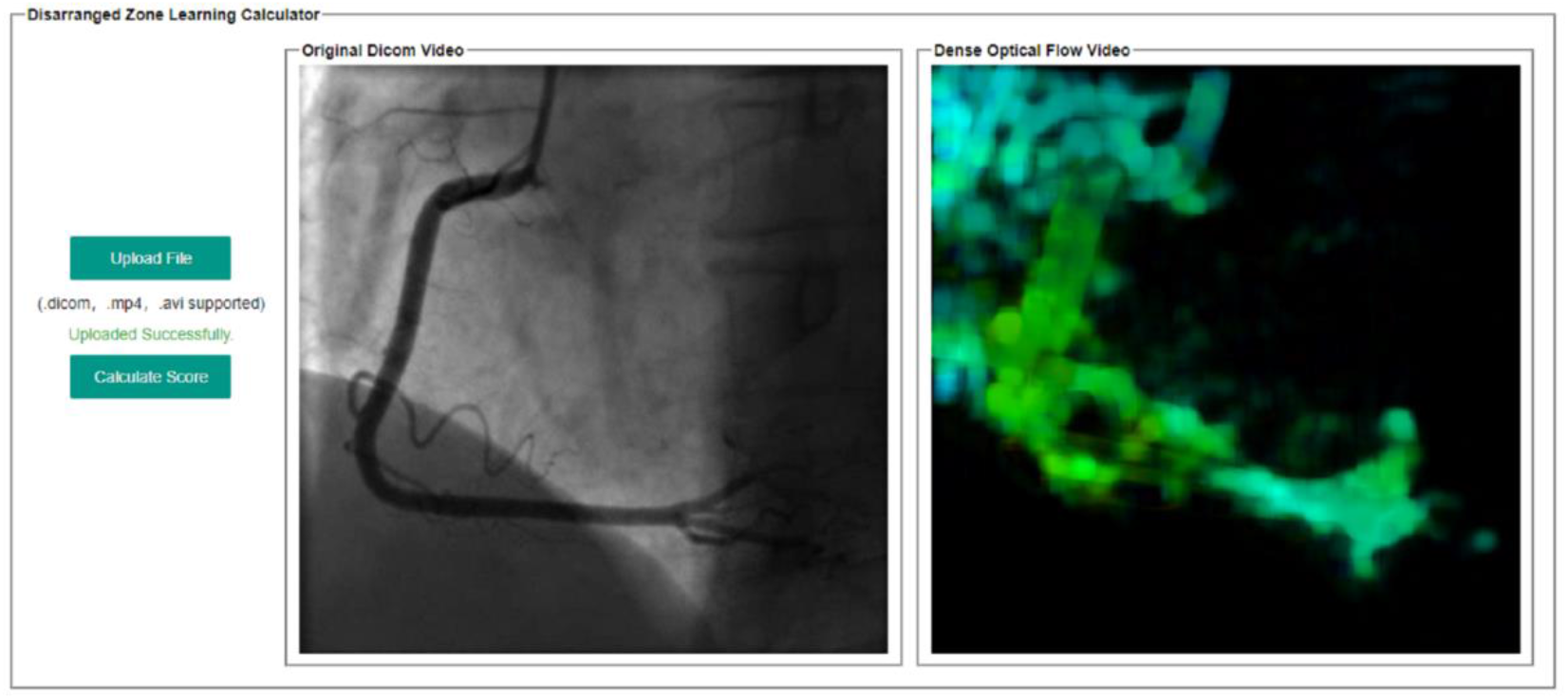
Software Interface.

**Supplemental Figure 8.**
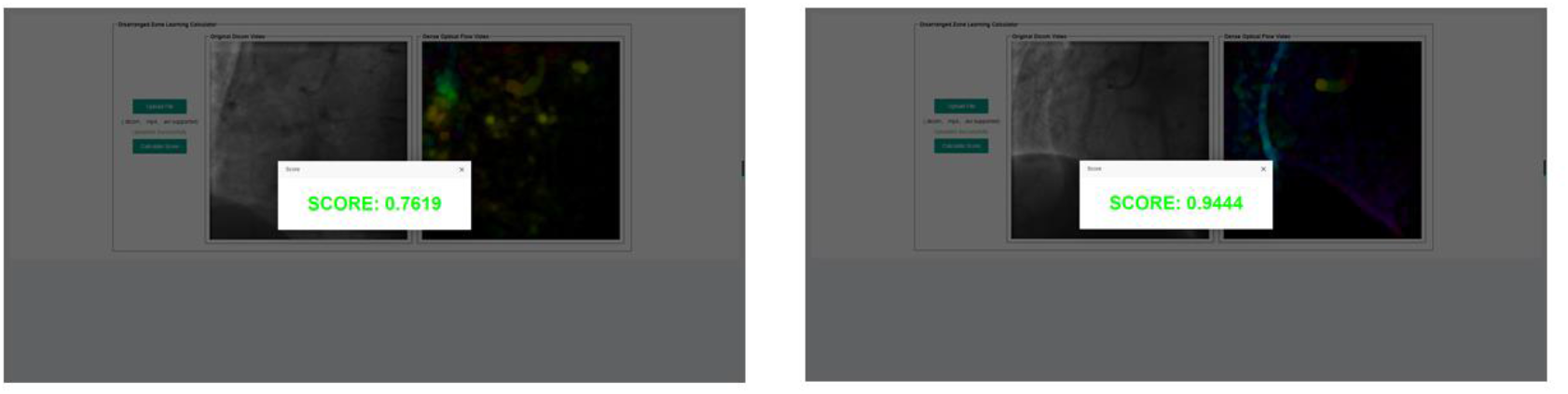
Scores Generation. The left RCA coronary angiography uploaded in our example is an abnormal video with occlusion while the right RCA coronary angiography uploaded in our example is a normal video. The final SCOREs are calculated. The occlusion video gets 0.76 and the normal video gets 0.94.

**Supplemental Table 1.** Cohen Kappa Confusion Matrix of 12 Ordinary People: Non-PCI Conduction after CAG.

| Non-PCI Conduction after CAG | 1 | 2 | 3 | 4 | 5 |
| --- | --- | --- | --- | --- | --- |
| 1 | 24 | 11 | 1 | 0 | 0 |
| 2 | 0 | 24 | 5 | 5 | 2 |
| 3 | 0 | 1 | 27 | 7 | 1 |
| 4 | 0 | 0 | 2 | 21 | 13 |
| 5 | 12 | 0 | 1 | 3 | 20 |

**Supplemental Table 2.** Cohen Kappa Confusion Matrix of 12 Ordinary People: PCI Conduction after CAG.

| PCI Conduction after CAG | 1 | 2 | 3 | 4 | 5 |
| --- | --- | --- | --- | --- | --- |
| 1 | 39 | 13 | 4 | 9 | 7 |
| 2 | 12 | 24 | 8 | 14 | 14 |
| 3 | 10 | 20 | 22 | 17 | 3 |
| 4 | 7 | 5 | 22 | 24 | 14 |
| 5 | 4 | 10 | 16 | 8 | 34 |

**Supplemental Table 3.** Cohen Kappa Confusion Matrix of 7 physicians: Non-PCI Conduction after CAG.

| Non-PCI Conduction after CAG | 1 | 2 | 3 | 4 | 5 |
| --- | --- | --- | --- | --- | --- |
| 1 | 20 | 1 | 0 | 0 | 0 |
| 2 | 0 | 19 | 2 | 0 | 0 |
| 3 | 0 | 1 | 18 | 2 | 0 |
| 4 | 0 | 0 | 1 | 19 | 1 |
| 5 | 1 | 0 | 0 | 0 | 20 |

**Supplemental Table 4.** Cohen Kappa Confusion Matrix of 7 physicians: PCI Conduction after CAG.

| PCI Conduction after CAG | 1 | 2 | 3 | 4 | 5 |
| --- | --- | --- | --- | --- | --- |
| 1 | 30 | 2 | 1 | 3 | 6 |
| 2 | 4 | 25 | 4 | 2 | 7 |
| 3 | 5 | 2 | 25 | 10 | 0 |
| 4 | 1 | 6 | 8 | 20 | 7 |
| 5 | 2 | 7 | 4 | 7 | 22 |

**Supplemental Table 5.**
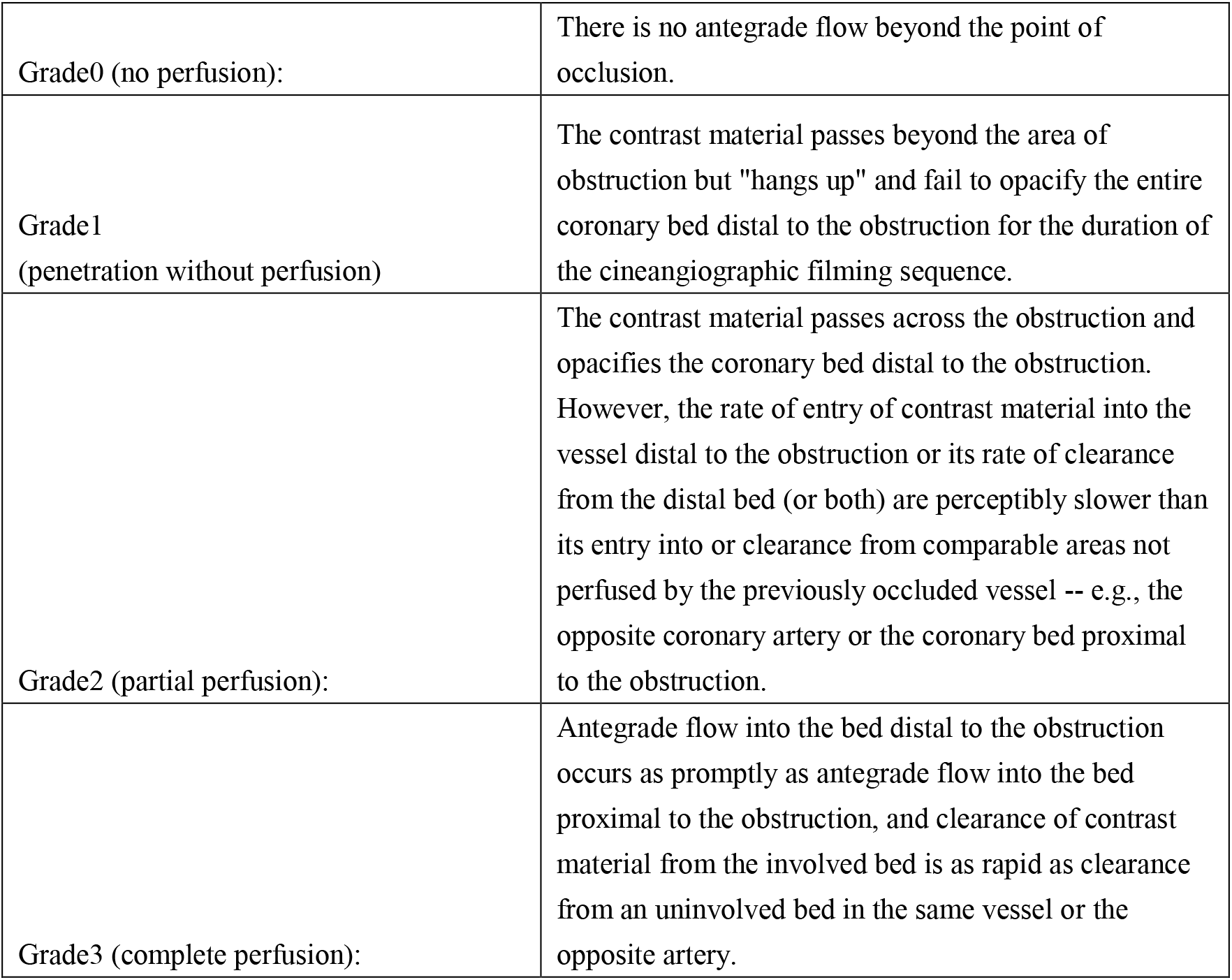
Definitions of Perfusion in the TIMI Trial.

## 5. The questionnaire

### Disordered Sequence Recovery Questionnaire

A normal coronary angiography displays a sequential antegrade contrast material development process in the coronary artery. The development status indicates the status and properties of the lesions. For stenosis or occluded vessels, the antegrade process will be hindered or even blocked.

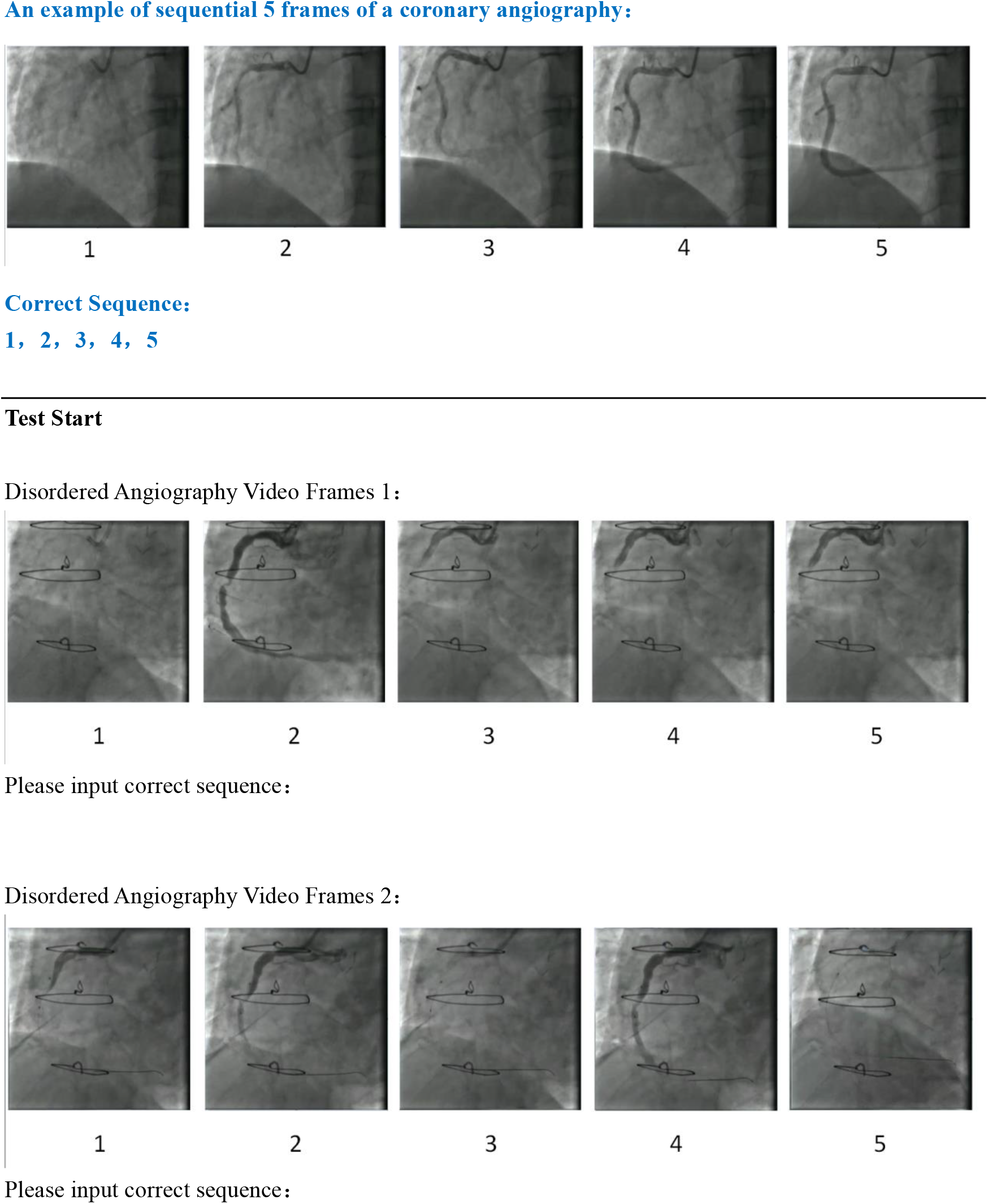
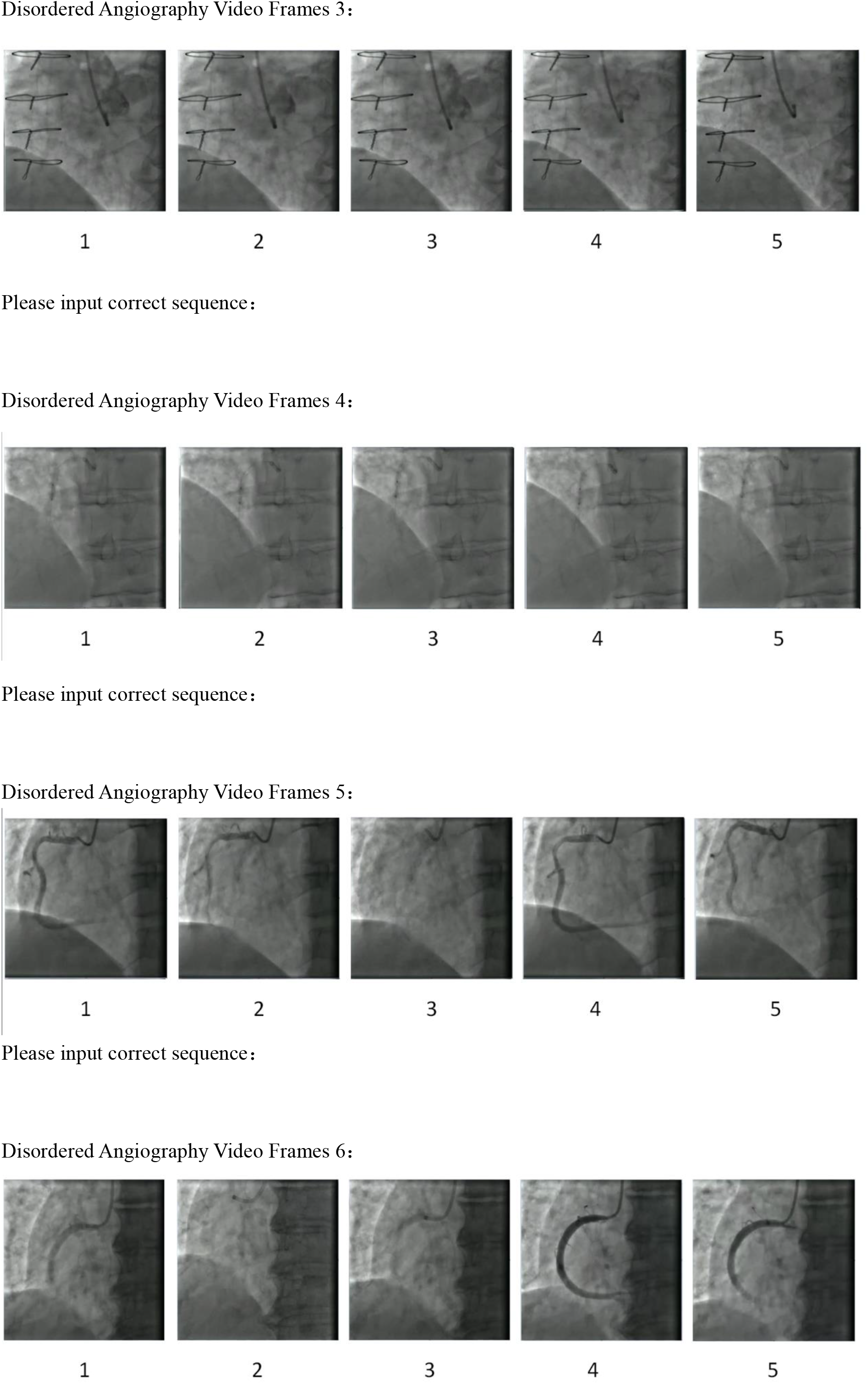
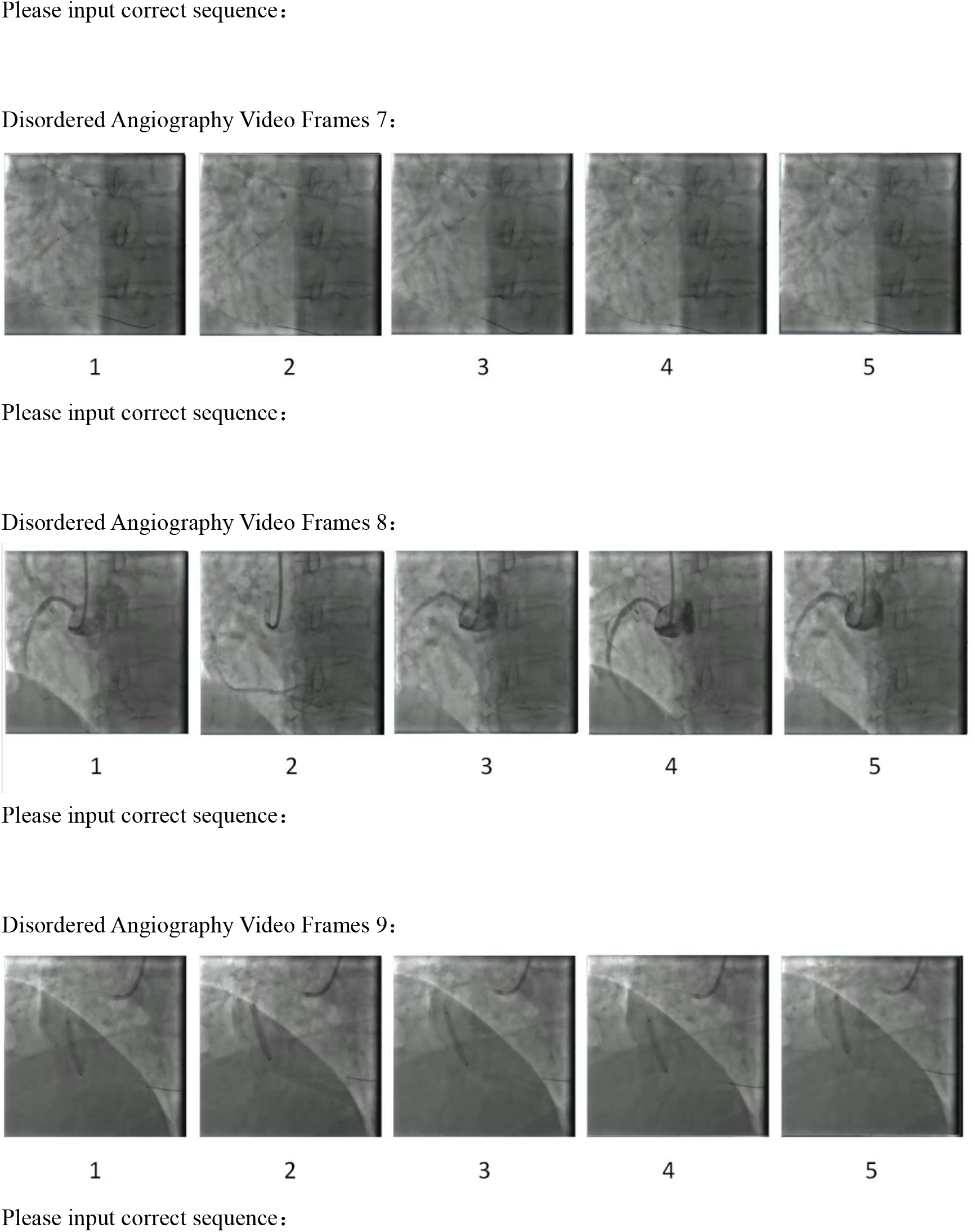

